# Factors Associated with Receipt and Parental Intent for COVID-19 Vaccination of Children Ages 5–11 years

**DOI:** 10.1101/2022.06.24.22276865

**Authors:** Tammy A. Santibanez, Jessica Penn Lendon, James A. Singleton, Carla L. Black, Tianyi Zhou, Jennifer L. Kriss, Anurag Jain, Laurie D. Elam-Evans, Nina B. Masters, Georgina Peacock

## Abstract

**Background and Objectives:** COVID-19 vaccine was first recommended for children ages 5–11 years on November 2, 2021. This report describes COVID-19 vaccination coverage and parental intent to vaccinate their child ages 5–11 years, overall, by sociodemographic characteristics, and by social and behavioral drivers of vaccination, the fourth month after recommendation.

**Methods:** We analyzed data from 5,438 interviews conducted in February 2022 from the National Immunization Survey-Child COVID Module (NIS-CCM), a national random-digit-dial cellular telephone survey of households with children.

**Results:** 30.9% of children ages 5–11 were vaccinated with ≥1 dose of COVID-19 vaccine, 35.2% were unvaccinated and the parent reported they probably or definitely would get the child vaccinated or were unsure, and 33.9% were unvaccinated and the parent probably or definitely would not get the child vaccinated. Vaccination coverage and parental intent differed by sociodemographic variables, including income, health insurance status, and rurality. Parental intent to vaccinate children also differed by ethnicity and race. Concern about the child getting COVID-19 and confidence in vaccine importance and safety were positively associated with vaccination receipt and intent to get the child vaccinated.

**Conclusions:** By the fourth month of the COVID-19 vaccination program for children ages 5–11 years, less than one-third were vaccinated, and coverage was lower for some sociodemographic subgroups. An additional one-third of children had a parent who was open to vaccinating the child. Efforts to address parental concerns regarding vaccine safety and to convey the importance of the vaccine might improve vaccination coverage.

## INTRODUCTION

COVID-19 is usually milder in children than adults; however, children can become severely ill and require hospitalization and intensive care, and some children have prolonged illness duration.^1,2^ Some children have also contracted multisystem inflammatory syndrome after infection with COVID-19.^3^ Children might play an important role in the transmission of COVID-19, highlighting the importance of child vaccinations in the effort to reduce COVID-19 community transmission and hospitalization rates.^4^ The U.S. Food and Drug Administration authorized the emergency use of the Pfizer-BioNTech COVID-19 vaccine for children ages 5–11 years in the United States on October 29, 2021, which was endorsed on November 2, 2021, by the Centers for Disease Control and Prevention (CDC) based on recommendations from the Advisory Committee on Immunization Practices.^5^ Administrative data reported to CDC showed that coverage among children 5–11 years was 24% during the first two months of vaccine rollout, which was lower than coverage among those ages 12–15 years (33%) during the first two months of eligibility for that age group.^6^ A report on the safety of the Pfizer-BioNTech COVID-19 vaccine, the only COVID-19 vaccine authorized for 5-17 year olds, after administration of approximately 8 million doses to children ages 5–11 years found the vaccine to be safe, with mild to moderate local and systemic reactions to be expected after vaccination.^7^ In two studies completed within 3–4 months of vaccine authorization, COVID-19 vaccine effectiveness after two doses among children ages 5–11 years was 31% against asymptomatic and symptomatic Omicron infection and 46% against COVID-19 associated emergency department and urgent care encounters.^8,9^

The evaluation of disparities in receipt of routine childhood vaccinations is an important component of the national immunization program in the United States.^10^ Understanding sociodemographic differences in COVID-19 vaccination and parental intent to vaccinate children ages 5–11 years during the first few months after the COVID-19 vaccine recommendation is essential in efforts to achieve higher vaccination coverage in this age group and addressing inequities in coverage early in the vaccination program. Better understanding of the associations between behavioral and social drivers of vaccine uptake and the willingness of parents to vaccinate their children might help to identify specific issues and demographic subgroups where tailored efforts might be helpful to improve vaccination coverage.

This report describes COVID-19 vaccination coverage among children ages 5–11 years during the first four months of eligibility and parents’ intent to vaccinate their children, overall and by sociodemographic characteristics using data from the National Immunization Survey-Child COVID Module (NIS-CCM). The study also examines variation in several behavioral and social drivers of COVID-19 vaccination by parents’ intent to vaccinate their children.

## METHODS

The NIS-CCM is a random-digit-dial cellular telephone survey of households with children ages 5–17 years. The NIS-CCM began on July 22, 2021, including households with children ages 13–17 years and asking the respondent about the child’s COVID-19 vaccination status and their intention to vaccinate the child. On October 1^st^, NIS-CCM was expanded to add children ages 12 and 5–11 years (intent questions only), and on November 2^nd^ vaccination status questions were added for children ages 5–11. The NIS-CCM utilizes the same sampling frame used for the NIS-Child.^11,12^ The respondent to a NIS survey is the parent or guardian of the child who self-reports as being knowledgeable about the child’s vaccination history (hereafter referred to as parent).

COVID-19 vaccination coverage was based on the parent’s response to the question “Has [child] received at least one dose of a COVID-19 vaccine?” Parental intent among parents who reported the child did not receive a vaccine was measured by asking: “How likely are you to get [child] a COVID-19 vaccine? Would you say you would: definitely get a vaccine for [child]; probably get a vaccine; probably not get a vaccine; definitely not get a vaccine; or are not sure?” We grouped children as follows: 1) child vaccinated with ≥1 dose, 2) “child unvaccinated and parent open to vaccination”, defined as unvaccinated children whose parent reported they definitely or probably will get the child vaccinated or were unsure, and 3) “child unvaccinated and parent reluctant to vaccinate”, defined as unvaccinated children whose parent reported they definitely or probably will not get the child vaccinated.

Further, in order to control for the effect that any increasing vaccination coverage in a group has in depleting the proportion with high intent in the pool of those still unvaccinated (because of them getting vaccinated over time), we created an additional variable called “willingness” to get the child vaccinated with COVID-19 vaccine. “Willingness” included parents who already had the child vaccinated (they are willing to get the child vaccinated, as evidenced by them doing so) and parents open to vaccination (i.e. parents with unvaccinated children who definitely or probably will get the child vaccinated or were unsure). Parents who reported that the child had received COVID-19 vaccination were asked the kind of place where the child received the vaccination. All information about the parent, child, and household was self-reported by the parent respondent.

The variables describing potential drivers of COVID-19 vaccine uptake are derived from the Behavioral and Social Drivers of Vaccination (BeSD) Framework.^13^ The domains and associated survey questions were:

᎐ *Thinking and Feeling*:
  - “How concerned are you about [child’s name] getting COVID-19?” “Not at all concerned, a little concerned, moderately concerned, or very concerned.”
  - “How important do you think getting a COVID-19 vaccine is to protect [child’s name] against COVID-19?” “Not at all important, a little important, somewhat important, or very important”
  - “How safe do you think a COVID-19 vaccine is for [child’s name]?” “Not at all safe, somewhat safe, very safe, or completely safe”.
᎐ *Social Processes*:
  - “If you had to guess, about how many of your family and friends have gotten a COVID-19 vaccine for their children ages 5–11 years?” “None, some, many, or almost all.”
  - “Has a doctor, nurse, or another health professional ever recommended that you get a COVID-19 vaccine for [child’s name]” “Yes or no.”
  - “Does [child’s name]’s school require a COVID-19 vaccine to attend in-person classes?” “Yes or no.”
᎐ *Practical Issues*:
  - “How difficult would it be/was it for you to get [child’s name] a COVID-19 vaccine?” “Not at all difficult, a little difficult, somewhat difficult, or very difficult.”

Data from interviews conducted from January 30 through February 26, 2022, were analyzed. There were 5,496 completed parent interviews for children ages 5–11 years. Of these, 35 were missing vaccination status, and 23 of the unvaccinated children did not have vaccination intent reported for the child, for a resulting analytic sample of n=5,438. The cumulative NIS-CCM response rate was 15.1%. Weighted proportions with 95% confidence intervals (CIs) were estimated considering the complex survey design and weights by using SAS-callable SUDAAN (version 11.0.3, RTI) and SAS version 9.4 (SAS Institute, Inc.). The NIS-CCM survey weights adjust for non-response and households without cellular telephones and were calibrated to match counts of children with one or more doses of COVID-19 vaccine by sex within each U.S. Department of Health and Human Services (HHS) region for children ages 5–11 years as of mid-month, using data reported to CDC by jurisdictions.^6^ T-tests for proportions were used to test for statistically significant differences with *p*<0.05. The NCHS reliability criteria for proportions was implemented and proportions not meeting the criteria were suppressed from the tables.^14^

## RESULTS

Figure 1 shows the distribution of children across six levels of vaccination and intent, including vaccinated with at least one dose, and each of the five levels of intent for unvaccinated children included in the survey question. COVID-19 vaccination coverage among children ages 5–11 years increased from 8.0% in November 2021 to 20.7% in December 2021, 27.6% in January 2022, and 30.9% in February 2022 (Figure 1). In February, 35.2% were unvaccinated and the parent reported they probably or definitely would get the child vaccinated or were unsure, and 33.9% were unvaccinated and the parent probably or definitely would not get the child vaccinated. The places that children ages 5–11 years received their vaccinations were: medical place (46.4% [specifically: 23.3% doctor’s office, 12.4% clinic/health center, 6.5% hospital, 3.4% health department, 0.9% other medical place]), pharmacy/drug store (38.3%), school (7.5%), mass vaccination site (5.3%), or another non-medical place (2.6%; data not shown).

**Figure 1.**
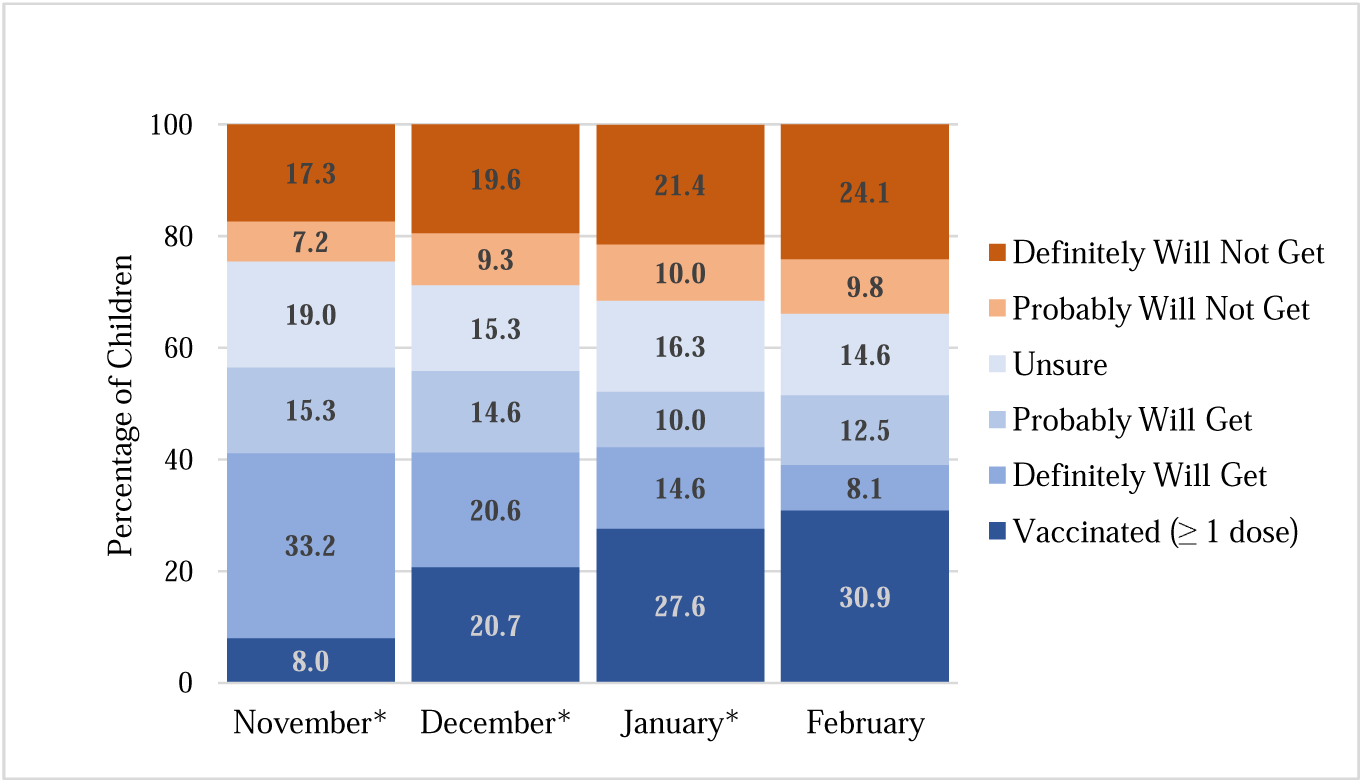
COVID-19 Vaccination Status (≥ 1 dose) and Parental Intent to Have the Child Vaccinated, by Month of Interview, Children Ages 5–11 Years, November 2021–February 2022, United States, National Immunization Survey-Child COVID Module (NIS-CCM) *Estimates from COVIDVaxView: https://www.cdc.gov/vaccines/imz-managers/coverage/covidvaxview/index.html

Child’s receipt of ≥1 dose of COVID-19 vaccine and parents’ willingness to get their child vaccinated with COVID-19 vaccine varied by sociodemographic characteristics (Table 1). A higher percentage of children ages 7–8 years (29.5%) and 9–11 years (34.4%) were vaccinated compared to children ages 5–6 years (26.0%), yet there were no age differences in parents’ willingness to get their child vaccinated (Table 1). There were no differences in the percentage of Hispanic or Black non-Hispanic children receiving ≥1 dose compared with White non-Hispanic children; however, there was a higher willingness to vaccinate among parents of Black non-Hispanic children (72.7%) and Hispanic children (71.4%) compared with White non-Hispanic children (57.9%, Table 1). The highest vaccination coverage (68.7%) and parental willingness to vaccinate (92.9%) was among parents of Asian non-Hispanic children.

**Table 1.**
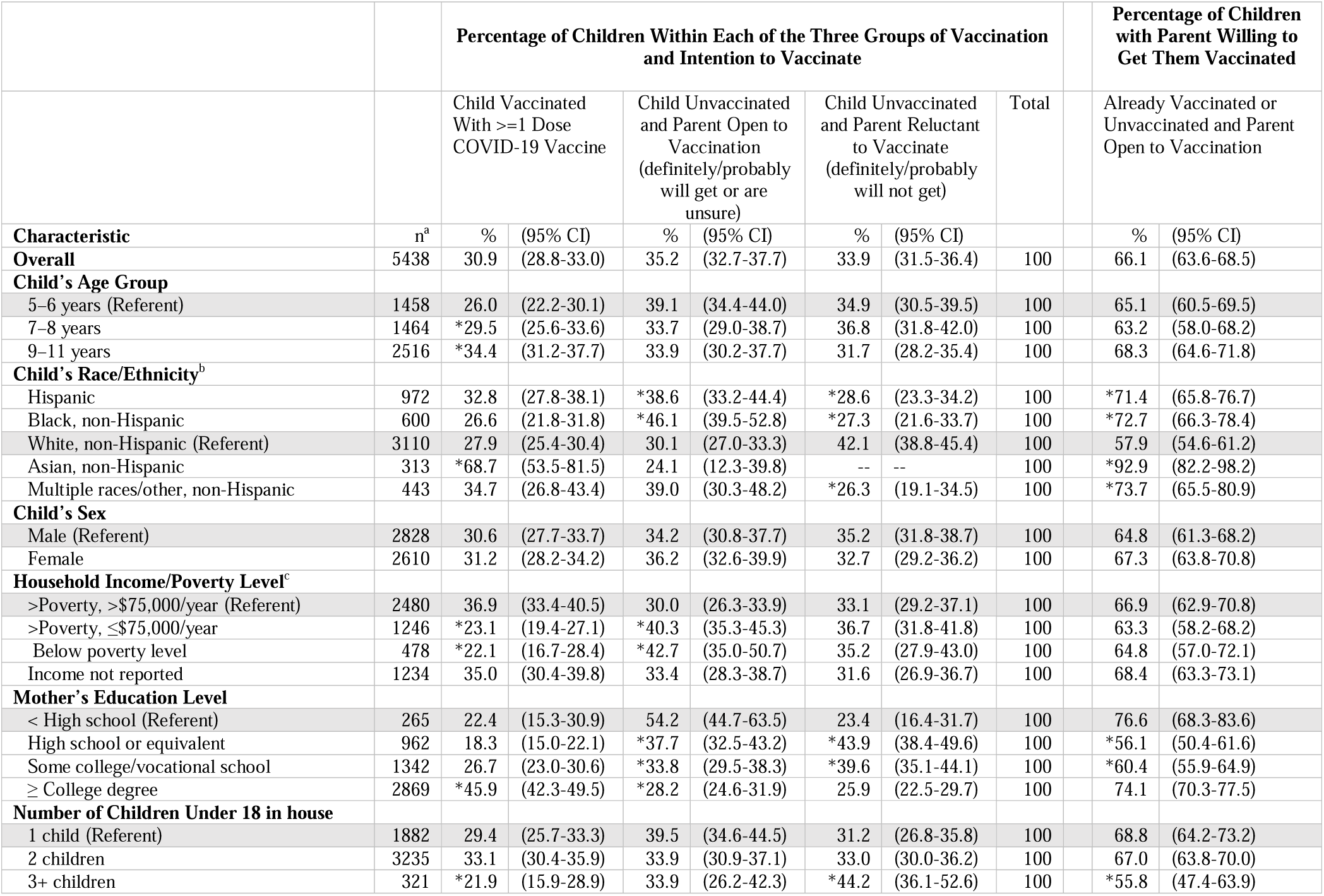
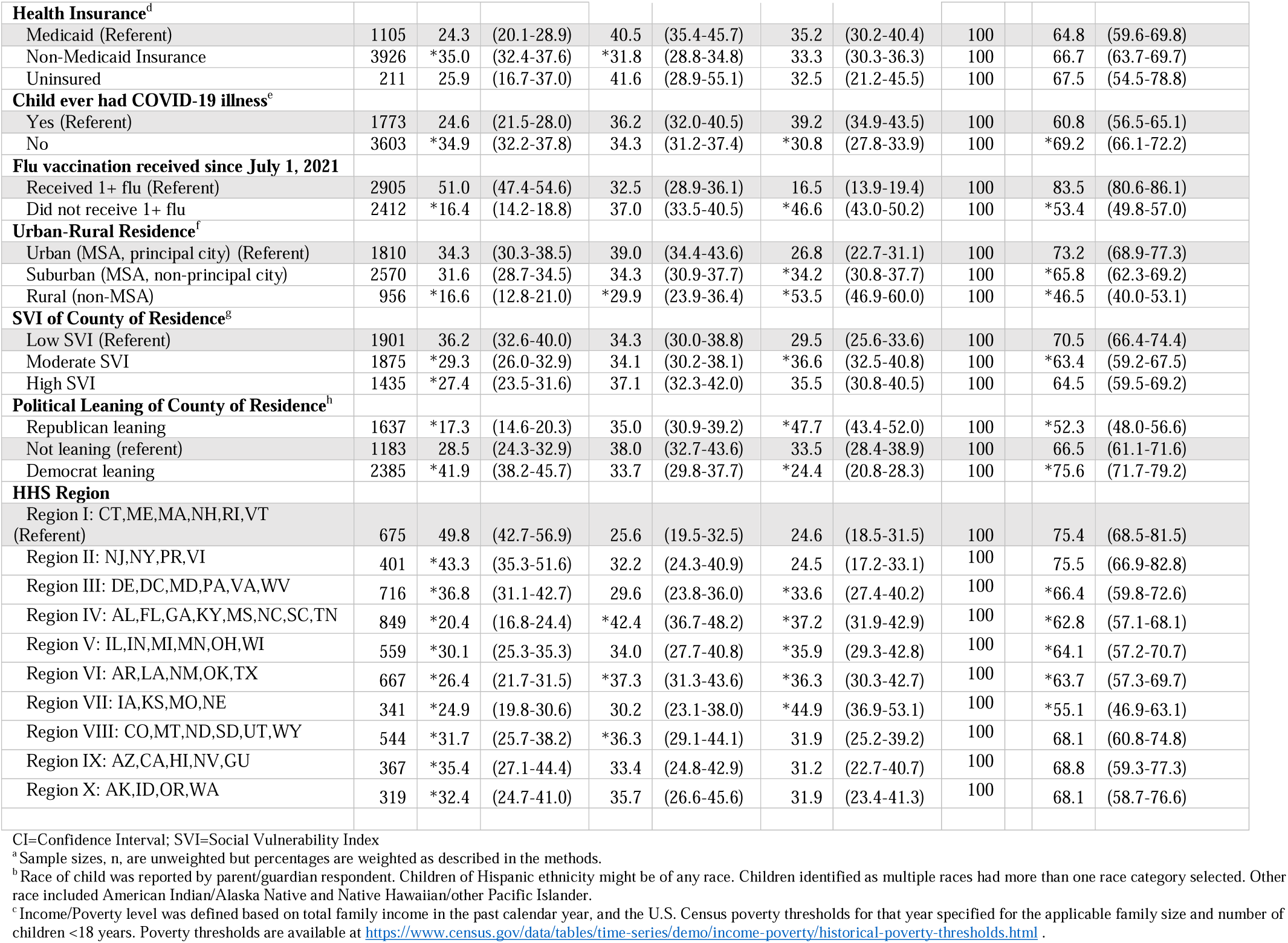
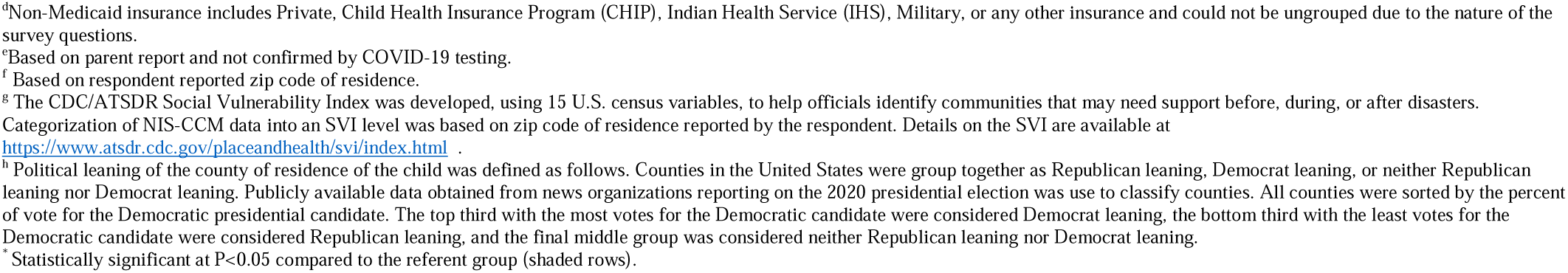
Sociodemographic Characteristics Associated with COVID-19 Vaccination Status and Intention of Parents to Vaccinate Children ages 5–11 years, February 2022, United States, National Immunization Survey-Child COVID Module (NIS-CCM)

A lower percentage of children received COVID-19 vaccination in households above poverty with income <=$75,000 (23.1%) and below the poverty level (22.1%) compared with children in households above poverty with income >$75,000 (36.9%); however, there was no difference in willingness to vaccinate by income (range 63-68% across income levels). A higher proportion of children with non-Medicaid health insurance received COVID-19 vaccination (35.0%) compared with Medicaid-insured children (24.3%), while uninsured children had similar coverage to Medicaid-insured children (25.9%); there were no differences in willingness to vaccinate by insurance status. Among children who did not receive an influenza vaccination since July 2021, 16.4% received a COVID-19 vaccine compared with 51.0% of children who had received an influenza vaccination. A lower proportion of children living in rural areas were vaccinated (16.6%) than children living in urban areas (34.3%), and there was a lower willingness to vaccinate in rural (46.5%) and suburban (65.8%) areas compared with urban areas (73.2%). There were also differences in vaccination coverage and willingness to vaccinate by county-level characteristics and by HHS region (see Table 1). The study also found variation by sociodemographic characteristics in parents’ openness and reluctance to vaccinate children who were unvaccinated, as shown in Table 1.

There were strong associations between social and behavioral drivers and the child’s receipt of ≥1 dose of COVID-19 vaccine and parental intent to vaccinate the child (Figure 2). Parents who were reluctant (reported that they definitely or probably would *not* get their child vaccinated) were less likely to report they were very/moderately concerned about the child getting COVID-19 when compared to parents of vaccinated children (23.6% vs 56.8%), were less likely to report that getting a COVID-19 vaccine was somewhat/very important (16.0% vs 95.9%), less likely to report that COVID-19 vaccine is completely/very safe (7.1% vs 80.0%), less likely to report that almost all/many friends/family have their child vaccinated (6.0% vs 63.6%), and less likely to report a provider recommendation for COVID-19 vaccine (18.4% vs 56.6%). Parents who were open to vaccinating their child, but had not yet vaccinated them (reported that they were definitely/probably/unsure), were less likely to report that getting a COVID-19 vaccine was somewhat/very important when compared to parents of vaccinated children (77.6% vs 95.9%), less likely to report that COVID-19 vaccine is completely/very safe (38.3% vs 80.0%), perception of fewer friends/family having their child vaccinated (16.0% vs 63.6%), and less likely to report a provider recommendation for COVID-19 vaccine (24.0% vs 56.6%).

**Figure 2.**
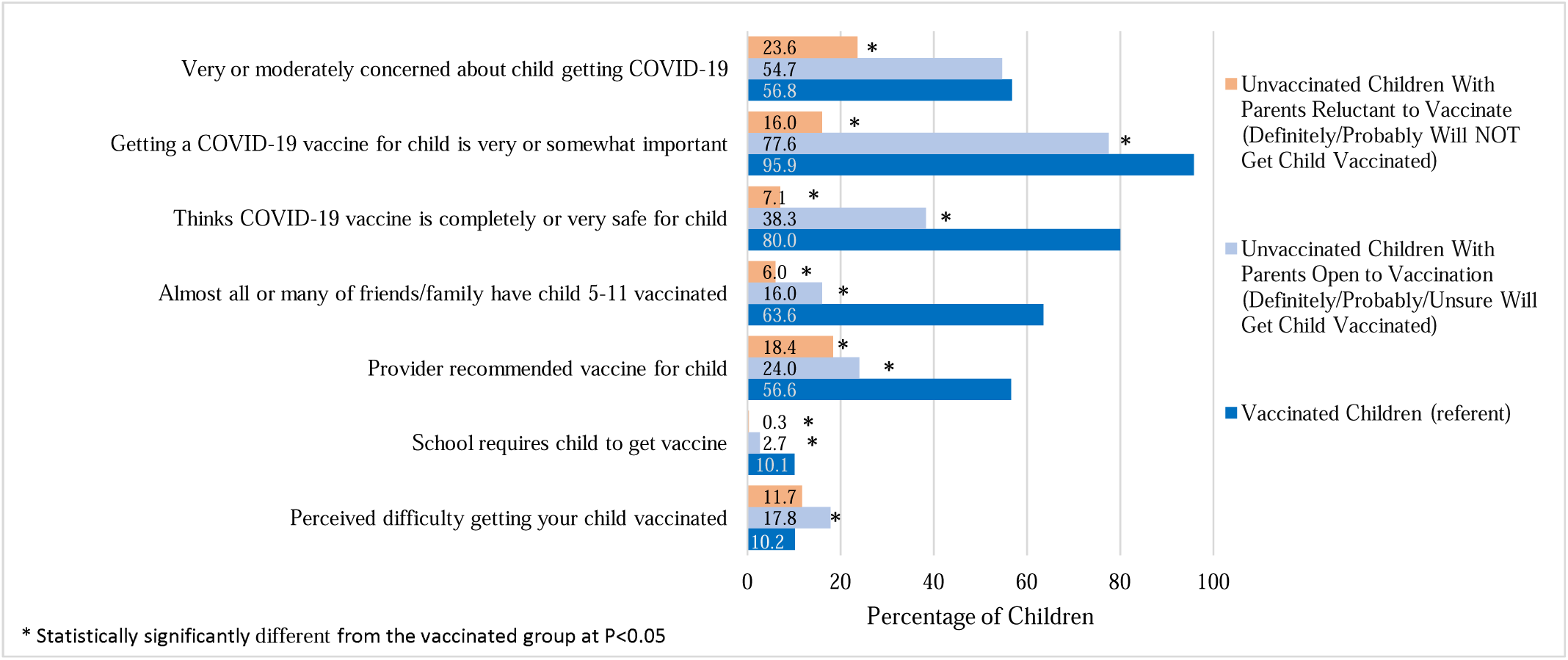
Behavioral and Social Drivers Associated with COVID-19 Vaccination, by Vaccination Status (≥ 1 Dose) and Intention of Parents to Vaccinate Children Ages 5–11 years, February 2022, United States, National Immunization Survey-Child COVID Module (NIS-CCM)

Table 2 describes the behavioral and social drivers for COVID-19 vaccination among the subset of unvaccinated children whose parents were open to vaccination. Hispanic children in this group had a higher percentage of parent-reported concern about the child getting COVID-19 (58.4%), but a lower percentage of confidence in the safety of the vaccine (29.7%) than among White non-Hispanic children (44.1% and 45.2%, respectively). Among unvaccinated Black non-Hispanic children whose parent was open to vaccination, there was a higher percentage of parent-reported concern about the child getting COVID-19 (67.8%) and perceived importance of the vaccine (88.2%) than among White non-Hispanic unvaccinated children (44.1% and 71.3%, respectively). For children who had a mother with a college degree, the respondents were more confident in the vaccine’s safety than those with ≤ high school education (50.4% vs 30.3%) and had higher reported provider recommendations for COVID-19 vaccination (30.7% vs 18.3%). Other sociodemographic differences are reported in Table 2.

**Table 2.**
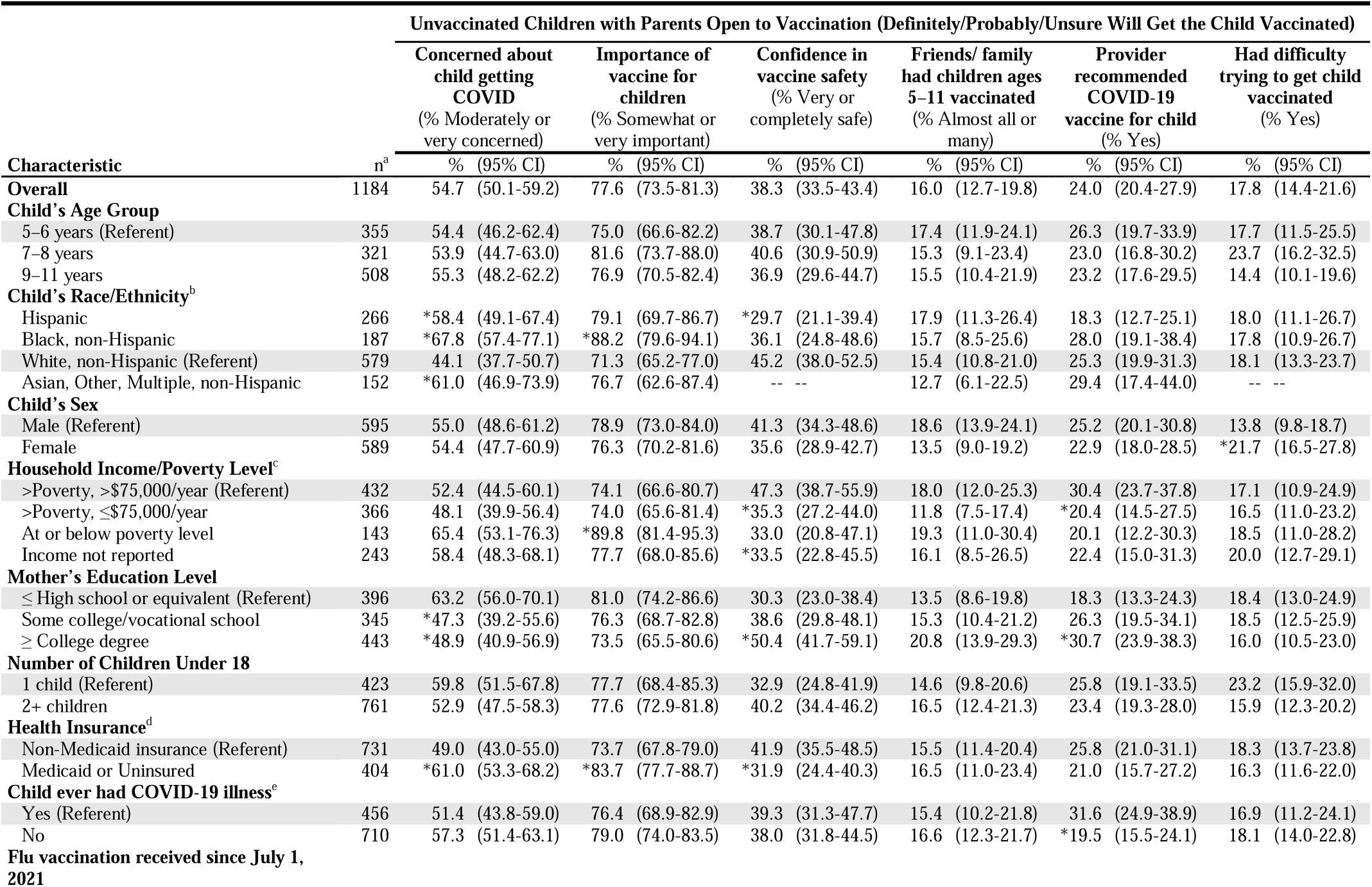
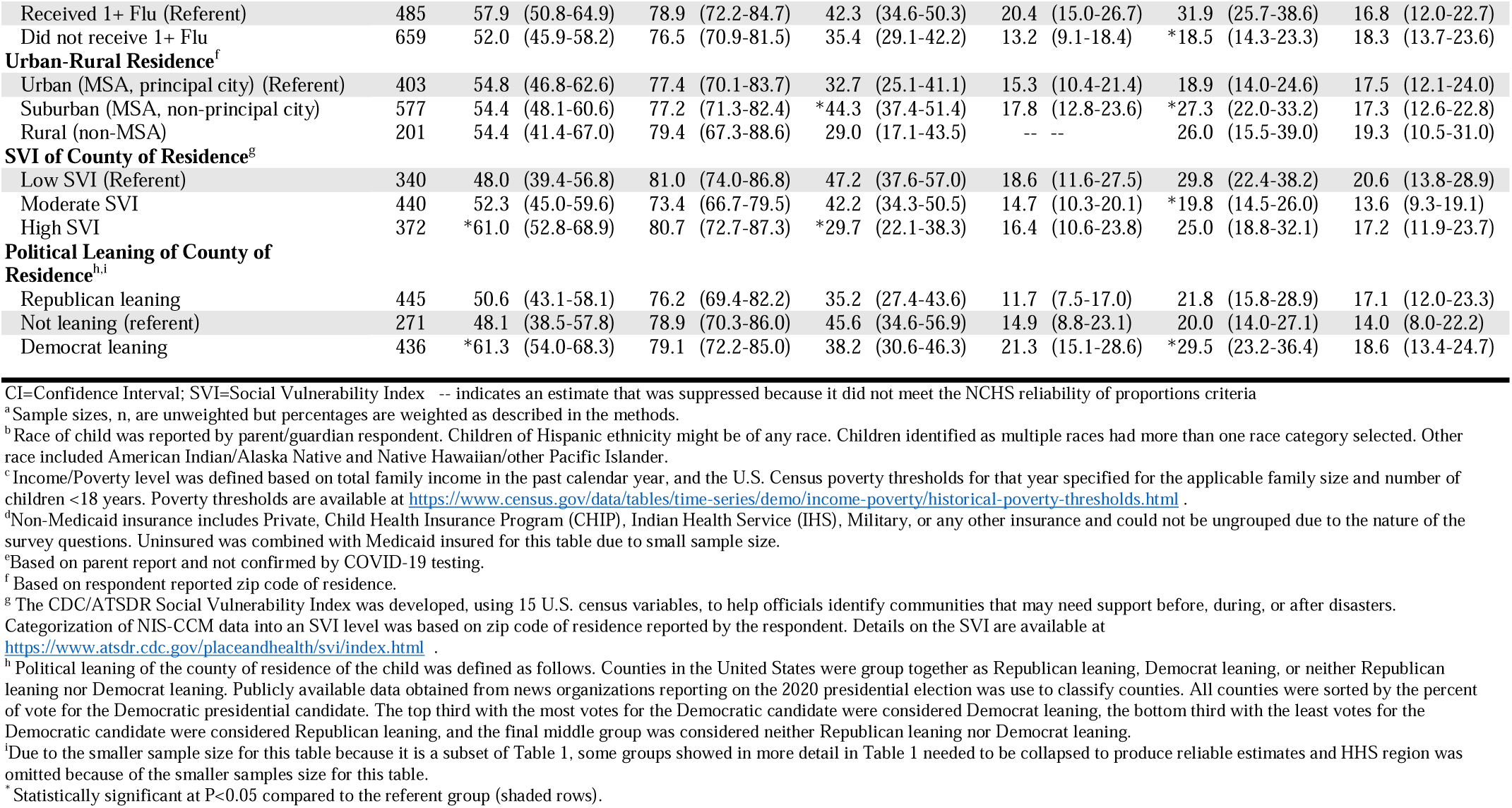
Sociodemographic Variation in Behavioral and Social Drivers for COVID-19 Vaccination, Among Unvaccinated Children Ages 5–11 years With Parents Who Definitely/Probably/Unsure Will Get the Child Vaccinated, United States, February 2022, National Immunization Survey-Child COVID Module (NIS-CCM)

| Characteristic | n<sup>a</sup> | Unvaccinated Children with Parents Open to Vaccination (Definitely/Probably/Unsure Will Get the Child Vaccinated) |  |  |  |  |  |  |
| --- | --- | --- | --- | --- | --- | --- | --- | --- |
|  |  | Concerned about<br>child getting<br>COVID<br>(% Moderately or<br>very concerned) | Importance of<br>vaccine for<br>children<br>(% Somewhat or<br>very important) | Confidence in<br>vaccine safety<br>(% Very or<br>completely safe) | Friends/ family<br>had children ages<br>5–11 vaccinated<br>(% Almost all or<br>many) | Provider<br>recommended<br>COVID-19<br>vaccine for child<br>(% Yes) | Had difficulty<br>trying to get child<br>vaccinated<br>(% Yes) |  |
|  |  | % (95% CI) | % (95% CI) | % (95% CI) | % (95% CI) | % (95% CI) | % (95% CI) |  |
| **Overall** | 1184 | 54.7 (50.1-59.2) | 77.6 (73.5-81.3) | 38.3 (33.5-43.4) | 16.0 (12.7-19.8) | 24.0 (20.4-27.9) | 17.8 (14.4-21.6) |  |
| **Child's Age Group** |  |  |  |  |  |  |  |  |
| 5–6 years (Referent) | 355 | 54.4 (46.2-62.4) | 75.0 (66.6-82.2) | 38.7 (30.1-47.8) | 17.4 (11.9-24.1) | 26.3 (19.7-33.9) | 17.7 (11.5-25.5) |  |
| 7–8 years | 321 | 53.9 (44.7-63.0) | 81.6 (73.7-88.0) | 40.6 (30.9-50.9) | 15.3 (9.1-23.4) | 23.0 (16.8-30.2) | 23.7 (16.2-32.5) |  |
| 9–11 years | 508 | 55.3 (48.2-62.2) | 76.9 (70.5-82.4) | 36.9 (29.6-44.7) | 15.5 (10.4-21.9) | 23.2 (17.6-29.5) | 14.4 (10.1-19.6) |  |
| **Child's Race/Ethnicity<sup>b</sup>** |  |  |  |  |  |  |  |  |
| Hispanic | 266 | *58.4 (49.1-67.4) | 79.1 (69.7-86.7) | *29.7 (21.1-39.4) | 17.9 (11.3-26.4) | 18.3 (12.7-25.1) | 18.0 (11.1-26.7) |  |
| Black, non-Hispanic | 187 | *67.8 (57.4-77.1) | *88.2 (79.6-94.1) | 36.1 (24.8-48.6) | 15.7 (8.5-25.6) | 28.0 (19.1-38.4) | 17.8 (10.9-26.7) |  |
| White, non-Hispanic (Referent) | 579 | 44.1 (37.7-50.7) | 71.3 (65.2-77.0) | 45.2 (38.0-52.5) | 15.4 (10.8-21.0) | 25.3 (19.9-31.3) | 18.1 (13.3-23.7) |  |
| Asian, Other, Multiple, non-Hispanic | 152 | *61.0 (46.9-73.9) | 76.7 (62.6-87.4) | -- -- | 12.7 (6.1-22.5) | 29.4 (17.4-44.0) | -- -- |  |
| **Child's Sex** |  |  |  |  |  |  |  |  |
| Male (Referent) | 595 | 55.0 (48.6-61.2) | 78.9 (73.0-84.0) | 41.3 (34.3-48.6) | 18.6 (13.9-24.1) | 25.2 (20.1-30.8) | 13.8 (9.8-18.7) |  |
| Female | 589 | 54.4 (47.7-60.9) | 76.3 (70.2-81.6) | 35.6 (28.9-42.7) | 13.5 (9.0-19.2) | 22.9 (18.0-28.5) | *21.7 (16.5-27.8) |  |
| **Household Income/Poverty Level<sup>c</sup>** |  |  |  |  |  |  |  |  |
| >Poverty, >\$75,000/year (Referent) | 432 | 52.4 (44.5-60.1) | 74.1 (66.6-80.7) | 47.3 (38.7-55.9) | 18.0 (12.0-25.3) | 30.4 (23.7-37.8) | 17.1 (10.9-24.9) | |
| >Poverty, ≤\$75,000/year | 366 | 48.1 (39.9-56.4) | 74.0 (65.6-81.4) | *35.3 (27.2-44.0) | 11.8 (7.5-17.4) | *20.4 (14.5-27.5) | 16.5 (11.0-23.2) | |
| At or below poverty level | 143 | 65.4 (53.1-76.3) | *89.8 (81.4-95.3) | 33.0 (20.8-47.1) | 19.3 (11.0-30.4) | 20.1 (12.2-30.3) | 18.5 (11.0-28.2) |  |
| Income not reported | 243 | 58.4 (48.3-68.1) | 77.7 (68.0-85.6) | *33.5 (22.8-45.5) | 16.1 (8.5-26.5) | 22.4 (15.0-31.3) | 20.0 (12.7-29.1) |  |
| **Mother's Education Level** |  |  |  |  |  |  |  |  |
| ≤ High school or equivalent (Referent) | 396 | 63.2 (56.0-70.1) | 81.0 (74.2-86.6) | 30.3 (23.0-38.4) | 13.5 (8.6-19.8) | 18.3 (13.3-24.3) | 18.4 (13.0-24.9) |  |
| Some college/vocational school | 345 | *47.3 (39.2-55.6) | 76.3 (68.7-82.8) | 38.6 (29.8-48.1) | 15.3 (10.4-21.2) | 26.3 (19.5-34.1) | 18.5 (12.5-25.9) |  |
| ≥ College degree | 443 | *48.9 (40.9-56.9) | 73.5 (65.5-80.6) | *50.4 (41.7-59.1) | 20.8 (13.9-29.3) | *30.7 (23.9-38.3) | 16.0 (10.5-23.0) |  |
| **Number of Children Under 18** |  |  |  |  |  |  |  |  |
| 1 child (Referent) | 423 | 59.8 (51.5-67.8) | 77.7 (68.4-85.3) | 32.9 (24.8-41.9) | 14.6 (9.8-20.6) | 25.8 (19.1-33.5) | 23.2 (15.9-32.0) |  |
| 2+ children | 761 | 52.9 (47.5-58.3) | 77.6 (72.9-81.8) | 40.2 (34.4-46.2) | 16.5 (12.4-21.3) | 23.4 (19.3-28.0) | 15.9 (12.3-20.2) |  |
| **Health Insurance<sup>d</sup>** |  |  |  |  |  |  |  |  |
| Non-Medicaid insurance (Referent) | 731 | 49.0 (43.0-55.0) | 73.7 (67.8-79.0) | 41.9 (35.5-48.5) | 15.5 (11.4-20.4) | 25.8 (21.0-31.1) | 18.3 (13.7-23.8) |  |
| Medicaid or Uninsured | 404 | *61.0 (53.3-68.2) | *83.7 (77.7-88.7) | *31.9 (24.4-40.3) | 16.5 (11.0-23.4) | 21.0 (15.7-27.2) | 16.3 (11.6-22.0) |  |
| **Child ever had COVID-19 illness<sup>e</sup>** |  |  |  |  |  |  |  |  |
| Yes (Referent) | 456 | 51.4 (43.8-59.0) | 76.4 (68.9-82.9) | 39.3 (31.3-47.7) | 15.4 (10.2-21.8) | 31.6 (24.9-38.9) | 16.9 (11.2-24.1) |  |
| No | 710 | 57.3 (51.4-63.1) | 79.0 (74.0-83.5) | 38.0 (31.8-44.5) | 16.6 (12.3-21.7) | *19.5 (15.5-24.1) | 18.1 (14.0-22.8) |  |
| **Flu vaccination received since July 1, 2021** |  |  |  |  |  |  |  |  |

|  |  |  |  |  |  |  |  |
| --- | --- | --- | --- | --- | --- | --- | --- |
| Received 1+ Flu (Referent) | 485 | 57.9 (50.8-64.9) | 78.9 (72.2-84.7) | 42.3 (34.6-50.3) | 20.4 (15.0-26.7) | 31.9 (25.7-38.6) | 16.8 (12.0-22.7) |
| Did not receive 1+ Flu | 659 | 52.0 (45.9-58.2) | 76.5 (70.9-81.5) | 35.4 (29.1-42.2) | 13.2 (9.1-18.4) | *18.5 (14.3-23.3) | 18.3 (13.7-23.6) |
| **Urban-Rural Residence<sup>f</sup>** |  |  |  |  |  |  |  |
| Urban (MSA, principal city) (Referent) | 403 | 54.8 (46.8-62.6) | 77.4 (70.1-83.7) | 32.7 (25.1-41.1) | 15.3 (10.4-21.4) | 18.9 (14.0-24.6) | 17.5 (12.1-24.0) |
| Suburban (MSA, non-principal city) | 577 | 54.4 (48.1-60.6) | 77.2 (71.3-82.4) | *44.3 (37.4-51.4) | 17.8 (12.8-23.6) | *27.3 (22.0-33.2) | 17.3 (12.6-22.8) |
| Rural (non-MSA) | 201 | 54.4 (41.4-67.0) | 79.4 (67.3-88.6) | 29.0 (17.1-43.5) | -- -- | 26.0 (15.5-39.0) | 19.3 (10.5-31.0) |
| **SVI of County of Residence<sup>g</sup>** |  |  |  |  |  |  |  |
| Low SVI (Referent) | 340 | 48.0 (39.4-56.8) | 81.0 (74.0-86.8) | 47.2 (37.6-57.0) | 18.6 (11.6-27.5) | 29.8 (22.4-38.2) | 20.6 (13.8-28.9) |
| Moderate SVI | 440 | 52.3 (45.0-59.6) | 73.4 (66.7-79.5) | 42.2 (34.3-50.5) | 14.7 (10.3-20.1) | *19.8 (14.5-26.0) | 13.6 (9.3-19.1) |
| High SVI | 372 | *61.0 (52.8-68.9) | 80.7 (72.7-87.3) | *29.7 (22.1-38.3) | 16.4 (10.6-23.8) | 25.0 (18.8-32.1) | 17.2 (11.9-23.7) |
| **Political Leaning of County of Residence<sup>h,i</sup>** |  |  |  |  |  |  |  |
| Republican leaning | 445 | 50.6 (43.1-58.1) | 76.2 (69.4-82.2) | 35.2 (27.4-43.6) | 11.7 (7.5-17.0) | 21.8 (15.8-28.9) | 17.1 (12.0-23.3) |
| Not leaning (referent) | 271 | 48.1 (38.5-57.8) | 78.9 (70.3-86.0) | 45.6 (34.6-56.9) | 14.9 (8.8-23.1) | 20.0 (14.0-27.1) | 14.0 (8.0-22.2) |
| Democrat leaning | 436 | *61.3 (54.0-68.3) | 79.1 (72.2-85.0) | 38.2 (30.6-46.3) | 21.3 (15.1-28.6) | *29.5 (23.2-36.4) | 18.6 (13.4-24.7) |

CI=Confidence Interval; SVI=Social Vulnerability Index -- indicates an estimate that was suppressed because it did not meet the NCHS reliability of proportions criteria
<sup>a</sup> Sample sizes, n, are unweighted but percentages are weighted as described in the methods.
<sup>b</sup> Race of child was reported by parent/guardian respondent. Children of Hispanic ethnicity might be of any race. Children identified as multiple races had more than one race category selected. Other race included American Indian/Alaska Native and Native Hawaiian/other Pacific Islander.
<sup>c</sup> Income/Poverty level was defined based on total family income in the past calendar year, and the U.S. Census poverty thresholds for that year specified for the applicable family size and number of children <18 years. Poverty thresholds are available at https://www.census.gov/data/tables/time-series/demo/income-poverty/historical-poverty-thresholds.html.
<sup>d</sup> Non-Medicaid insurance includes Private, Child Health Insurance Program (CHIP), Indian Health Service (IHS), Military, or any other insurance and could not be ungrouped due to the nature of the survey questions. Uninsured was combined with Medicaid insured for this table due to small sample size.
<sup>e</sup> Based on parent report and not confirmed by COVID-19 testing.
<sup>f</sup> Based on respondent reported zip code of residence.
<sup>g</sup> The CDC/ATSDR Social Vulnerability Index was developed, using 15 U.S. census variables, to help officials identify communities that may need support before, during, or after disasters.
Categorization of NIS-CCM data into an SVI level was based on zip code of residence reported by the respondent. Details on the SVI are available at

## DISCUSSION

This study found that less than one-third of children ages 5–11 years had received a COVID-19 vaccination by the fourth month after the vaccine was recommended for this age group. The lower uptake for this age group was similarly reported in other studies.^6,15^ We found that an additional 35.2% of children had parents who were open to vaccination. These parents could potentially be encouraged to vaccinate their child if vaccine confidence was increased, and access issues were reduced or removed.^16,17^ The 33.9% of children with parents who were reluctant to get their child vaccinated are not likely to get vaccinated without a larger shift in the parents’ beliefs.^18^

This study found no differences in COVID-19 vaccination coverage between Hispanic or Black non-Hispanic children compared with White non-Hispanic children. This absence of racial/ethnic disparity is encouraging and might be a result of the efforts at outreach to communities to promote and increase access to vaccination.^15,17^ Estimates available earlier in the vaccination program showed there might have been some racial/ethnic differences, but they had narrowed by February 2022 when the data for this study were collected.^6,19^ It is less encouraging that COVID-19 vaccination coverage was low (< 40%) for all racial/ethnic groups, except for Asian non-Hispanic children. The higher COVID-19 vaccination coverage among Asian non-Hispanic children is consistent with other studies and with what has been found for influenza vaccination.^6,20^

We did find disparities by household income and by health insurance status. Children in lower-income families had COVID-19 vaccination coverage that was approximately 15 percentage points lower than those in a higher-income household, yet the parents in these lower-income households were as willing to have their children vaccinated as were those in the higher-income households. This indicates possible structural access barriers to COVID-19 vaccination among lower-income children. Likewise, we found that children who were Medicaid-insured or uninsured had COVID-19 vaccination coverage that was approximately 10 percentage points lower than children with private or other non-Medicaid health insurance, with no differences in parental willingness to get the child vaccinated by insurance coverage. This disparity could be associated with the difficulty of enrolling Vaccines for Children (VFC) providers, who serve children that are uninsured or Medicaid-insured, to stock and administer COVID-19 vaccines. Given changes in coverage of administration fees for vaccination of uninsured people that took effect on April 5, 2022, these disparities might become wider in the future, perhaps more so for children without a medical home; however, only a small proportion of children are uninsured.^21^ Any interventions to reduce access barriers to COVID-19 vaccination for young children, such as vaccinations offered in school settings or increasing the availability of vaccinators during convenient hours for working parents, should be encouraged.^17^

The behavioral and social driver variables that were most different between children already vaccinated compared to unvaccinated children whose parent was open to getting them vaccinated were concerns about vaccine safety, perceptions of how many friends and family were getting their children vaccinated, and provider recommendations for COVID-19 vaccination. The lower confidence in vaccine safety was even more prevalent among Hispanic parents. Discussions with trusted messengers, such as the child’s pediatrician or community and faith-based leaders, might help allay parents’ concerns.^22^ This might also increase parental perception that the social norm is to have children vaccinated with all CDC-recommended childhood vaccines, even if this is not true within their immediate circle of friends and families. The finding of a lower percentage of provider recommendations among unvaccinated children compared to vaccinated children suggests that outreach to pediatricians and other medical providers for young children might be needed to encourage them to use every opportunity to strongly recommended COVID-19 vaccination for their patients, as is recommended for other childhood vaccinations. This is especially important for VFC providers who are serving Medicaid and uninsured children. Children without a medical home are likely missing out on the opportunity to receive a provider recommendation, emphasizing the importance of all children having a medical home. Furthermore, providers that not only recommend vaccines for their patients but also offer vaccines during the visit have been shown to result in higher vaccination coverage levels.^23^ Improving the availability of COVID-19 vaccines at provider offices would help increase coverage. Other evidence-based approaches for providers include integrating vaccination into standard medical practice and sending reminders to parents; given less than 10% of children in this study were reported to be vaccinated at school, expanded school-located vaccination programs could be considered.^17^

This study is subject to at least four limitations. First, the response rates were low (15%) and bias in estimates might remain after weighting. Survey weights were therefore calibrated to the COVID-19 vaccine administration data by region, age group, and sex to mitigate possible bias from incomplete sample frame and nonresponse. Second, vaccination status was parent-reported and subject to recall bias and social desirability bias. However, recall bias of the child’s vaccination status might be reduced given the recency of the event (<4 months). While calibration to the COVID-19 vaccine administration data does not correct for misclassification of vaccination status on an individual level, it does mitigate impact of this misclassification on estimates of vaccination coverage. Third, we defined the shorthand term “willingness” to vaccinate the child to include parents who already had the child vaccinated plus those whose child was unvaccinated but they said they definitely or probably or were unsure they would get the child vaccinated, however these reported intentions might not translate to actual willingness to have the child vaccinated. Semantic arguments might also be presented for including the already vaccinated as willing, however other shorthand words/phrases had their own limitations. Finally, the survey is cross-sectional, so cause and effect relationships cannot be determined.

## CONCLUSION

By the fourth month of the COVID-19 vaccination program for children ages 5–11 years, slightly less than one-third were vaccinated and coverage was even lower for some sociodemographic subgroups. An additional one-third of children had a parent who was open to vaccination for the child. There were variations in vaccination coverage and parental intent by several sociodemographic characteristics and in the potential drivers of vaccination. Efforts to address parental concerns regarding vaccine safety and to convey the importance of the vaccine might improve vaccination coverage. This study also suggests health-care provider recommendation of COVID-19 vaccines and discussions with trusted messengers might also improve coverage among children ages 5-11 years. ^17^

## Data Availability

The data used in this study are not released as public use datasets but can be accessed within a National Center for Health Statistics (NCHS) Research Data Center.

https://www.cdc.gov/vaccines/imz-managers/nis/index.html

## DISCLAIMER

The findings and conclusions in this report are those of the authors and do not necessarily represent the official position of the Centers for Disease Control and Prevention. The use of trade names and commercial sources is for identification only and does not imply endorsement by the U.S. Department of Health and Human Services.

## Notes

### Competing Interest Statement

The authors have declared no competing interest.

### Funding Statement

This study did not receive any funding.

### Author Declarations

The IRBs of both the Centers for Disease Control and Prevention (CDC) and NORC at the University of Chicago gave approval for conduct of the National Immunization Survey-Child COVID-19 Module used for this study.

## REFERENCES

1. Kim L, Whitaker M, O’Halloran A, et al. Hospitalization Rates and Characteristics of Children Aged <18 Years Hospitalized with Laboratory-Confirmed COVID-19 - COVID-NET, 14 States, March 1-July 25, 2020. MMWR Morbidity and mortality weekly report. Aug 14 2020;69(32):1081–1088. doi:10.15585/mmwr.mm6932e3

2. Molteni E, Sudre CH, Canas LS, et al. Illness duration and symptom profile in symptomatic UK school-aged children tested for SARS-CoV-2. The Lancet Child & adolescent health. Oct 2021;5(10):708–718. doi:10.1016/s2352-4642(21)00198-x

3. CDC. Multisystem Inflammatory Syndrome in Children (MIS-C) Associated with Coronavirus Disease 2019 (COVID-19). Accessed 3/7/2022, https://emergency.cdc.gov/han/2020/han00432.asp

4. Szablewski CM, Chang KT, Brown MM, et al. SARS-CoV-2 Transmission and Infection Among Attendees of an Overnight Camp - Georgia, June 2020. MMWR Morbidity and mortality weekly report. Aug 7 2020;69(31):1023–1025. doi:10.15585/mmwr.mm6931e1

5. Woodworth KR, Moulia D, Collins JP, et al. The Advisory Committee on Immunization Practices’ Interim Recommendation for Use of Pfizer-BioNTech COVID-19 Vaccine in Children Aged 5-11 Years - United States, November 2021. MMWR Morbidity and mortality weekly report. Nov 12 2021;70(45):1579–1583. doi:10.15585/mmwr.mm7045e1

6. Murthy NC, Zell E, Fast HE, et al. Disparities in First Dose COVID-19 Vaccination Coverage among Children 5-11 Years of Age, United States. Emerg Infect Dis. Feb 28 2022;28(5)doi:10.3201/eid2805.220166

7. Hause AM, Baggs J, Marquez P, et al. COVID-19 Vaccine Safety in Children Aged 5-11 Years - United States, November 3-December 19, 2021. MMWR Morbidity and mortality weekly report. Dec 31 2021;70(5152):1755–1760. doi:10.15585/mmwr.mm705152a1

8. Fowlkes AL, Yoon SK, Lutrick K, et al. Effectiveness of 2-Dose BNT162b2 (Pfizer BioNTech) mRNA Vaccine in Preventing SARS-CoV-2 Infection Among Children Aged 5-11 Years and Adolescents Aged 12-15 Years - PROTECT Cohort, July 2021-February 2022. MMWR Morbidity and mortality weekly report. Mar 18 2022;71(11):422–428. doi:10.15585/mmwr.mm7111e1

9. Klein NP, Stockwell MS, Demarco M, et al. Effectiveness of COVID-19 Pfizer-BioNTech BNT162b2 mRNA Vaccination in Preventing COVID-19-Associated Emergency Department and Urgent Care Encounters and Hospitalizations Among Nonimmunocompromised Children and Adolescents Aged 5-17 Years - VISION Network, 10 States, April 2021-January 2022. MMWR Morbidity and mortality weekly report. Mar 4 2022;71(9):352–358. doi:10.15585/mmwr.mm7109e3

10. Roper L, Hall MAK, Cohn A. Overview of the United States’ Immunization Program. The Journal of infectious diseases. Sep 30 2021;224(12 Suppl 2):S443-s451. doi:10.1093/infdis/jiab310

11. Wolter KK, Smith PJ, Khare M, et al. Statistical Methodology of the National Immunization Survey, 2005-2014. Vital and health statistics Ser 1, Programs and collection procedures. Dec 2017;(61):1–107.

12. CDC. About the National Immunization Surveys (NIS). Accessed 3/7/2022, https://www.cdc.gov/vaccines/imz-managers/nis/about.html

13. Wiley KE, Levy D, Shapiro GK, et al. A user-centered approach to developing a new tool measuring the behavioural and social drivers of vaccination. Vaccine. Oct 8 2021;39(42):6283–6290. doi:10.1016/j.vaccine.2021.09.007

14. Parker JD, Talih M, Malec DJ, et al. National Center for Health Statistics Data Presentation Standards for Proportions. Vital and health statistics Series 2, Data evaluation and methods research. Aug 2017;(175):1-22.

15. Kim C, Yee R, Bhatkoti R, et al. COVID-19 Vaccine Provider Access and Vaccination Coverage Among Children Aged 5-11 Years - United States, November 2021-January 2022. MMWR Morbidity and mortality weekly report. Mar 11 2022;71(10):378–383. doi:10.15585/mmwr.mm7110a4

16. Shen SC, Dubey V. Addressing vaccine hesitancy: Clinical guidance for primary care physicians working with parents. Canadian family physician Medecin de famille canadien. Mar 2019;65(3):175–181.

17. CDC. 12 COVID-19 Vaccination Strategies for Your Community. Accessed 4/1/2022, “www.cdc.gov/vaccines/covid-19/vaccinate-with-confidence/community.html

18. Witteman HO. Addressing Vaccine Hesitancy With Values. Pediatrics. Aug 2015;136(2):215–7. doi:10.1542/peds.2015-0949

19. CDC. COVIDVaxView. Accessed 4/1/2022, https://www.cdc.gov/vaccines/imz-managers/coverage/covidvaxview/index.html

20. CDC. Flu Vaccination Coverage, United States, 2020–21 Influenza Season. Accessed 3/13/2022, https://www.cdc.gov/flu/fluvaxview/coverage-2021estimates.htm

21. Administration HHRS. HRSA COVID-19 Uninsured Program Claims Submission Deadline FAQs. Accessed 4/1/2022, https://www.hrsa.gov/coviduninsuredclaim/submission-deadline

22. Osakwe ZT, Osborne JC, Osakwe N, Stefancic A. Facilitators of COVID-19 vaccine acceptance among Black and Hispanic individuals in New York: A qualitative study. American journal of infection control. 2022/03/01/ 2022;50(3):268–272. doi:https://doi.org/10.1016/j.ajic.2021.11.004

23. Ding H, Black CL, Ball S, et al. Influenza Vaccination Coverage Among Pregnant Women - United States, 2016-17 Influenza Season. MMWR Morbidity and mortality weekly report. Sep 29 2017;66(38):1016–1022. doi:10.15585/mmwr.mm6638a2

